# Clinical characteristics and Outcomes of 500 patients with COVID Pneumonia – Results from a Single center (Southend University Hospital)

**DOI:** 10.1101/2020.08.13.20163030

**Authors:** Gouri Koduri, Sriya Gokaraju, Maria Darda, Vinod Warrier, Irina Duta, Fiona Hayes, Iman El Sayed, Yasser Ahmed

## Abstract

**Objectives:** To characterise the clinical features of hospitalised COVID-19 patients in a single centre during the first epidemic wave and explore potential predictive variables associated with outcomes such as mortality and the need for mechanical ventilation, using baseline clinical parameters.

**Methodology:** We conducted a retrospective review of electronic records for demographic, clinical and laboratory data, imaging and outcomes for 500 hospitalised patients between February 20^th^ and May 7^th^ 2020 from Southend University Hospital, Essex, UK. Multivariate logistic regression models were used to identify risk factors relevant to outcome.

**Results:** The mean age of the cohort admitted to hospital with Covid-19, was 69.4 and 290 (58%) were over 70. The majority were Caucasians, 437 (87%) with ≤2 comorbidities 280(56%). Most common were hypertension 186(37%), Cardiovascular disease 178(36%) and Diabetes 128 (26%), represented in a larger proportion on the mortality group. Mean CFS was 4 with Non - Survivors had significantly higher CFS 5 vs 3 in survivors, p<0.001. In addition, Mean CRP was significantly higher 150 vs 90, p<0.001 in Non-Survivors. We observed the baseline predictors for mortality were age, CFS and CRP.

**Conclusions:** In this single centre study, older and frailer patients with more comorbidities and a higher baseline CRP and creatinine were risk factors for worse outcomes. Integrated frailty and age-based risk stratification are essential, in addition to monitoring SFR (Sp02/Fi02) and inflammatory markers throughout the disease course to allow for early intervention to improve patient outcomes.

## Introduction

The COVID-19 pandemic is one of the worst infectious disease outbreaks of recent times; with the first wave in the UK, we have encountered 312,000 confirmed cases and 44,819 fatalities ^1^. The pandemic coronavirus disease 19 (COVID-19) is characterized by a highly variable course. While most patients experience only mild symptoms, a relevant proportion of patients develop severe disease progression up to respiratory failure. Several factors and mechanisms are proposed to influence COVID-19 pathogenesis. The most notable risk factor is age, followed by co-morbidities, including diabetes, obesity, cardiovascular and cerebrovascular diseases ^2-5^.

The mortality rate is variable; this is because of differences in the population demographics, the ascending curve, the method used to register COVID-19, and the health services ^6^. A recent report showed that mortality rate was 5.6% for China and 15.2% outside of China ^7^. Belgium has relatively high case fatality rates (16.34%), followed by France (15.65%), UK (14.21%), Italy (14.15%), Hungary (13.07%), Netherlands (12.91%), Sweden (12.21%) and USA (5.95%). The mortality excess has been primarily seen in the age group of ≥65 years globally with higher case fatality rates(CFR) in older patients with comorbidities ^8,9^

We have analysed clinical characteristics and prognostic factors that may be pertinent to patient outcomes, describing measures that have not been extensively described in existing literature such as the Clinical Frailty Scale (CFS), an efficient tool for assessing frailty, since this may be significant in determining outcomes for older patients ^10^. We hope that by adding to this growing body of evidence identifying potential predictive variables in outcomes - we can assist early intervention in these patients in order to prevent rapid clinical deterioration and offer medications that have shown evidence in improving outcomes ^11,12^.

## Methodology

In this retrospective study, 500 patients with COVID-19 infection who were admitted to Southend University Hospital from 20^th^ February to 7^th^ May 2020 were enrolled. Demographic variables collected were age, sex, and ethnicity. Clinical signs and symptoms included categorical and continuous variables, baseline vitals and symptoms. Imaging results comprised chest radiography (CXR) abnormality and computed tomographic (CT) imaging. SP02 to Fi02 ratio (SFR) was calculated in addition to baseline partial arterial oxygen pressure (Pao2). Laboratory findings comprised full blood count, neutrophil to lymphocyte ratio (NLR), C - reactive protein and renal function.

A number of comorbidities and comorbidities of particular interest including pulmonary disease, diabetes, hypertension, coronary artery disease, cerebrovascular disease, cancer and chronic renal disease were recorded. We also collected data on degree of frailty, using the Rockwood Clinical Frailty Scale (CFS) on all patients, outcomes, total length of stay (LOS) and need for mechanical ventilation. We analysed the demographic, clinical, laboratory and imaging features of 500 patients with COVID-19 to determine potential biomarkers that may affect the prognosis of these patients.

## Statistical analysis

The baseline characteristics of all enrolled patients in survivor and non-survivor groups were summarized and compared by applying Student’s *t* test, the Chi-square test, and the Mann-Whitney *U* test as appropriate. We did not calculate sample size prior to conducting our study. However, based on a rule of thumb, we achieved a minimum required sample size for the development of the model based on the need for 10-15 non-survived patients per risk factor ^13^.

Quantitative data were described by mean (standard deviation) and median (minimum-maximum). Categorical variables were summarized by frequency and percent. Bivariate analysis using Independent sample t-test, Mann-Whitney test as well as Pearson’s Chi-square test compared different demographic and clinical parameters between survivors and non-survivors. Statistically significant and clinically relevant predictors were fitted in multivariate stepwise backward logistic regression analysis. Variables initially included were age, gender, CFS, Comorbidities >2, NLR, CRP, creatinine, RR, CPAP, SFR, Total LOS and Interaction CPAP*CFS. Model selection was judged by goodness of the fit using Likelihood Ratio Test as well as pseuso R^2^. Model cross-validation was performed by randomly splitting the sample into development and test sets (ratio 3:1). The prognostic ability of the model was determined by calculating the accuracy of model’s predicted probability as well as the area under the receiver operating characteristics curve (AUROC) on the test set. Statistical analysis was performed using IBM SPSS statistics program and R software packages “caTools”, “Imtest”, “caret”, “ROCR” and” ggplot2”. All statistical tests were two-sided and judged at 0.05 significance level.

## Results

The demographic and clinical parameters of the cohort, Survivors and Non survivors are shown in **Table 1**.

**Table 1:**
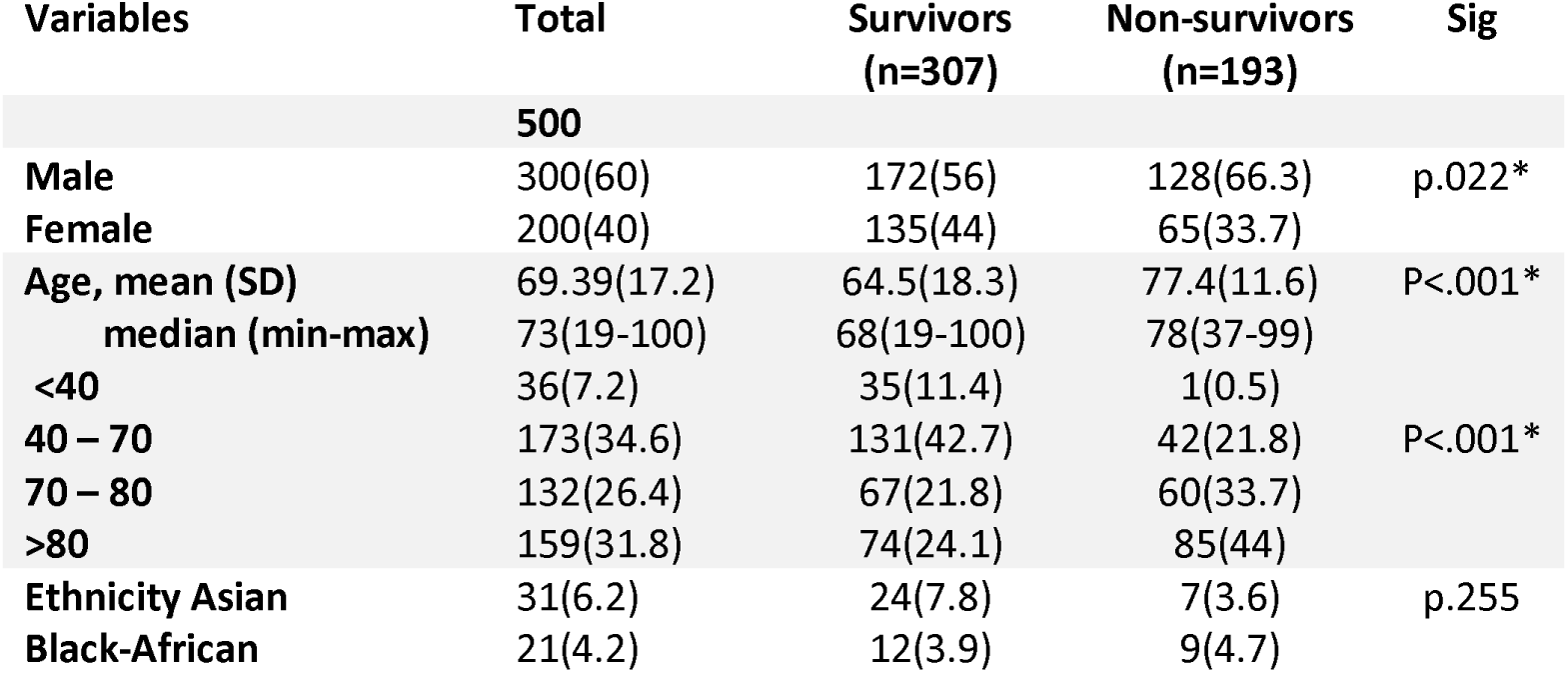
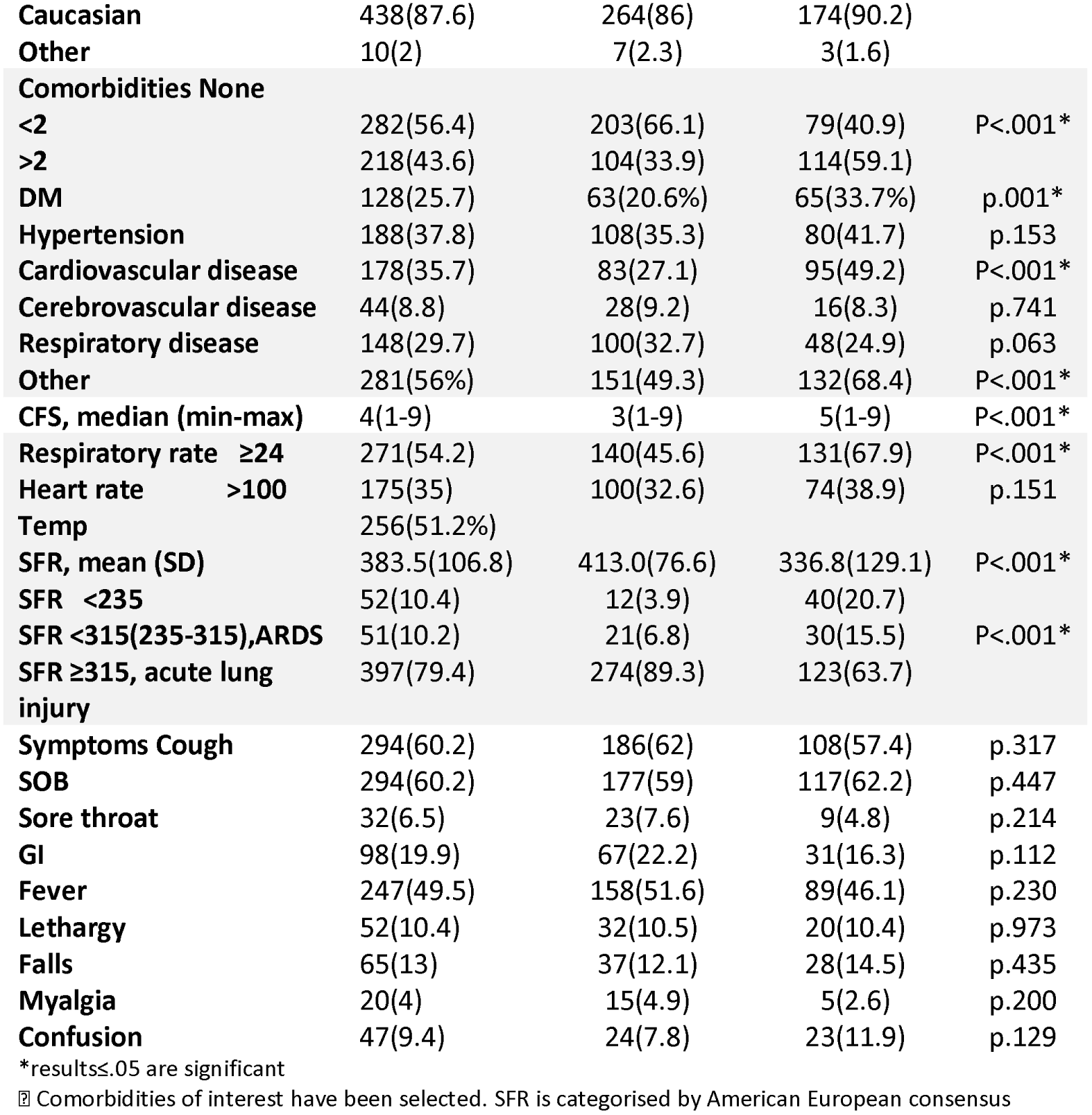
Cohort Characteristics.

Cough and dyspnoea were the most common presentation with equal representation 294(60.2%) followed by fever 247(49.5%), GIT symptoms 98(19.9%), falls 65(13%) and confusion 47(9.4%). Falls and confusion were common in elderly patients. Majority of the cohort was Caucasian, 438(87.6%) and age > 70 were 291 (58.9%).

Of the 500 patients 193(38.6%) died. There was male preponderance among Non - Survivors 128(66.3%) and were much older (77.4 vs 64.5 years, *P* < 0.001) and presented with more comorbidities, including Diabetes (65 [33.7%] vs. 63 [20.6%], *P* = 0.001 and Cardiovascular disease (95 [49.2%] vs. 83 [27.1%], *P* < 0.001). Proportion of deaths with PaO_2_/FiO_2_ less than 336 (mean) was statistically significantly (p < 0.001). As per ARDS criteria, Non-Survivors had lower SFR< 315, p< 0.001. Non-Survivors were more tachypnoeic, Respiratory rate > 24, p<0.001.

Clinical Frailty Scale: Mean CFS was 4, However compared to survivors ofCOVID-19, Non-Survivors had significantly higher CFS 3 vs 5, p<0.001.

A number of laboratory parameters showed significant differences among Survivors and Non-Survivors.

**Table 2**. Mean CRP was significantly higher 150 vs 90, p<0.001 in Non-Survivors, as well as Neutrophil count 7.84, p<0.001, Urea 12.71, p< 0.001 and creatinine 136, p=0.001.CXR abnormalities were observed more in Non survivors and supplemental Oxygen requirement was higher in NonSurvivors 181(93.8%) as compared to Survivors 165(53.9%), p< 0.001.

**Table 2:**
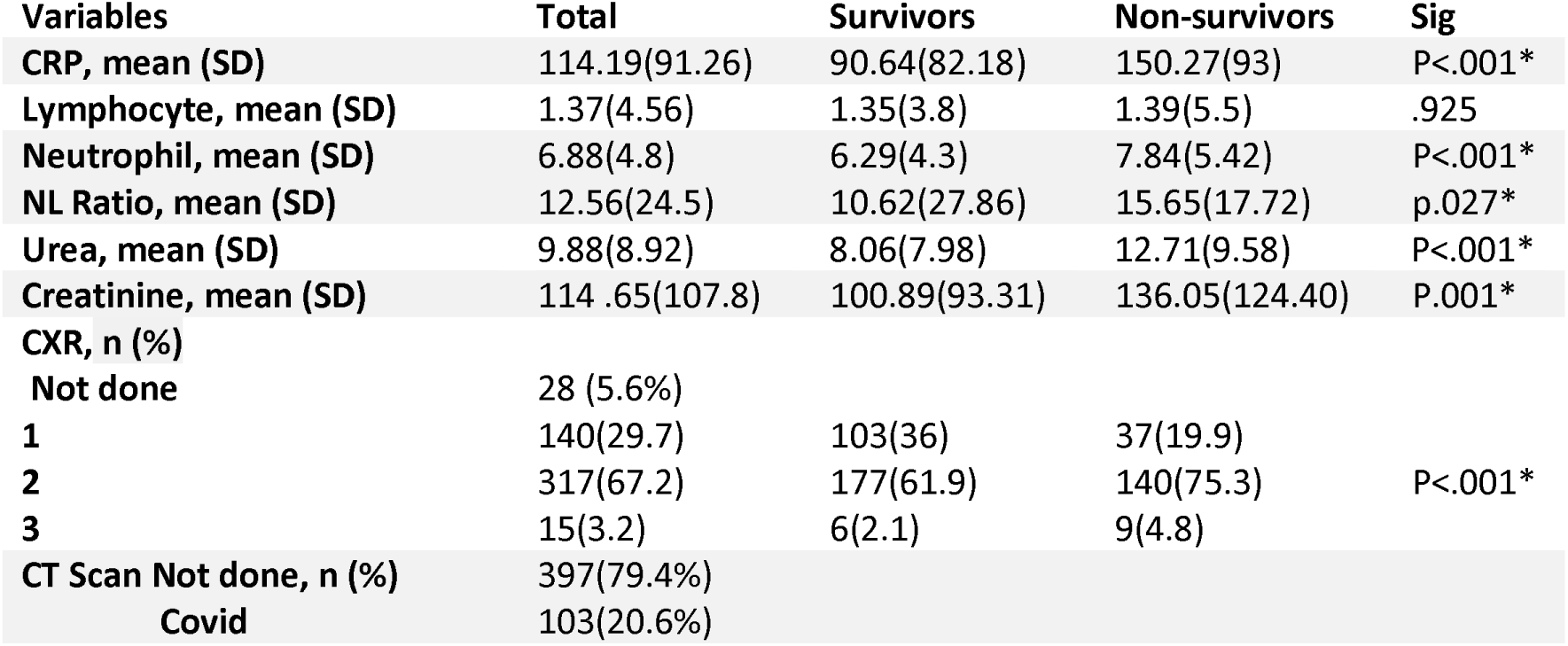
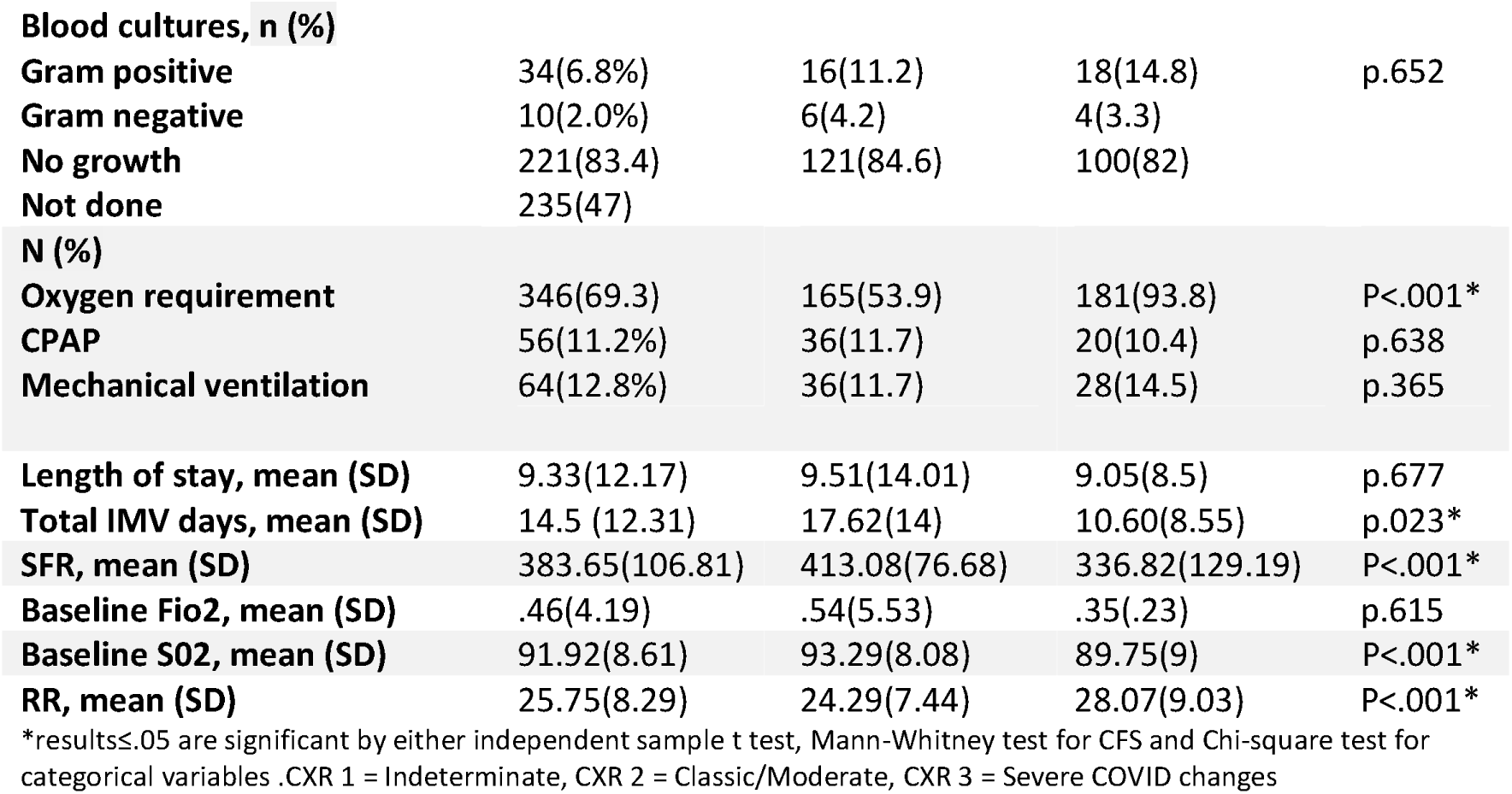
Baseline Clinical parameters.

There were no statistical differences on length of stay, need for mechanical ventilation or symptoms between the two groups.

### Predictors for Mortality, Multivariate Analysis

Next, we examined the variables which showed a significant correlation with negative outcomes in the multivariate logistic regression models to identify if these were independent predictors for mortality.

In the stepwise logistic regression models, the following were independent risk factors for mortality in model 1, age adjusted OR 1.035(95% CI 1.012 - 1.058), NLR adjusted OR 1.021 (95% CI 1.00 - 1.04), CFS adjusted OR 1.132(95% CI 1.13 -1.53) and CRP adjusted OR 1.006(95% CI 1.003 - 1.009), **(Table 3 and figure 1a)**. Again in model 2, age and CFS score were strong risk factors. Interestingly gender didn’t reach the statistical significance for mortality. Similarly creatinine, SFR and CXR abnormalities did not reach statistical significance but had a trend towards increased mortality **(Table 3 and fig 1b)**.

**Table 3:**
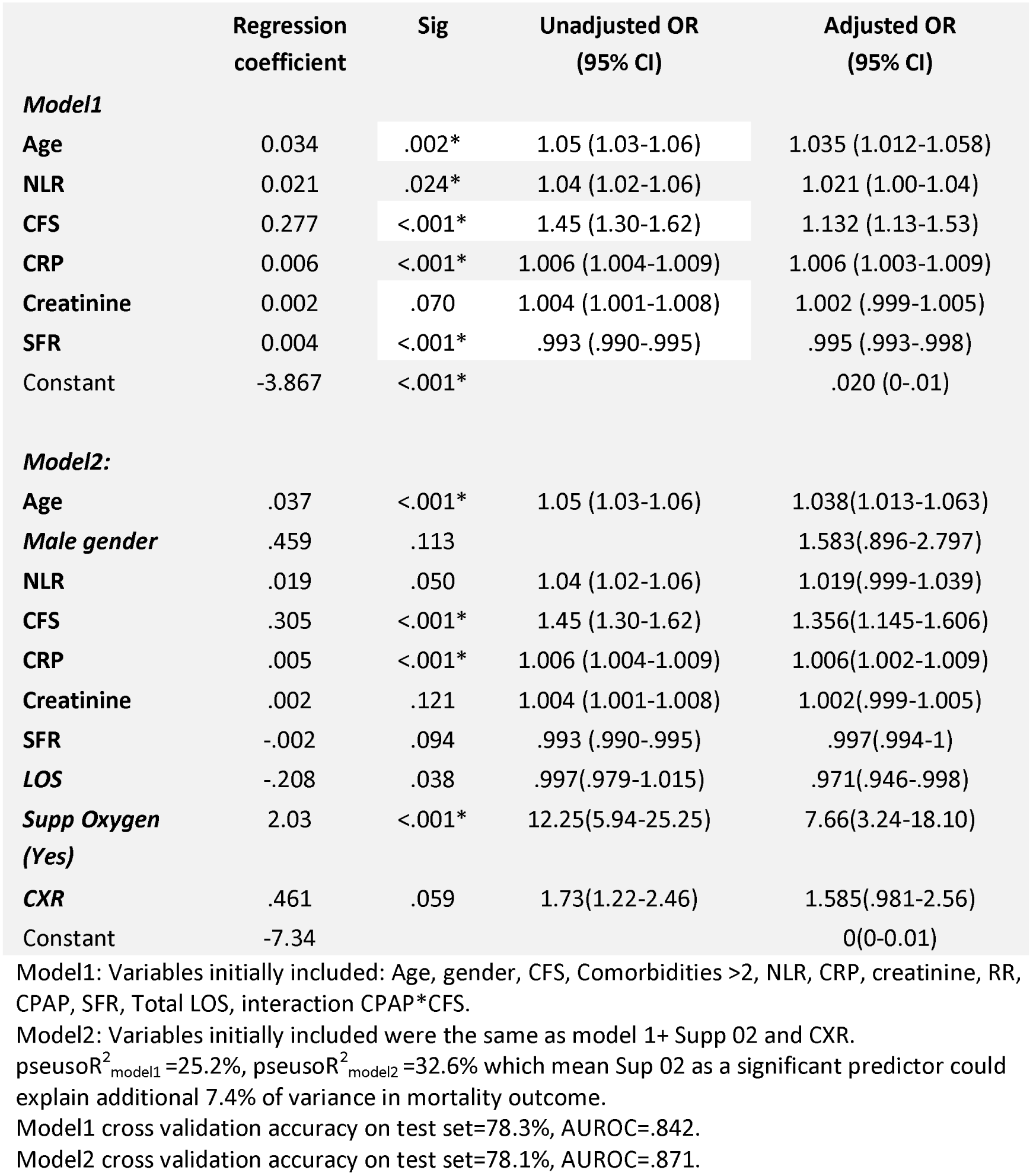
Multivariate logistic regression analysis for assessing independent predictors for mortality.

|  | Regression coefficient | Sig | Unadjusted OR (95% CI) | Adjusted OR (95% CI) |
| --- | --- | --- | --- | --- |
| **Model1** |  |  |  |  |
| Age | 0.034 | .002* | 1.05 (1.03-1.06) | 1.035 (1.012-1.058) |
| NLR | 0.021 | .024* | 1.04 (1.02-1.06) | 1.021 (1.00-1.04) |
| CFS | 0.277 | <.001* | 1.45 (1.30-1.62) | 1.132 (1.13-1.53) |
| CRP | 0.006 | <.001* | 1.006 (1.004-1.009) | 1.006 (1.003-1.009) |
| Creatinine | 0.002 | .070 | 1.004 (1.001-1.008) | 1.002 (.999-1.005) |
| SFR | 0.004 | <.001* | .993 (.990-.995) | .995 (.993-.998) |
| Constant | -3.867 | <.001* |  | .020 (0-.01) |
| **Model2:** |  |  |  |  |
| Age | .037 | <.001* | 1.05 (1.03-1.06) | 1.038(1.013-1.063) |
| Male gender | .459 | .113 |  | 1.583(.896-2.797) |
| NLR | .019 | .050 | 1.04 (1.02-1.06) | 1.019(.999-1.039) |
| CFS | .305 | <.001* | 1.45 (1.30-1.62) | 1.356(1.145-1.606) |
| CRP | .005 | <.001* | 1.006 (1.004-1.009) | 1.006(1.002-1.009) |
| Creatinine | .002 | .121 | 1.004 (1.001-1.008) | 1.002(.999-1.005) |
| SFR | -.002 | .094 | .993 (.990-.995) | .997(.994-1) |
| LOS | -.208 | .038 | .997(.979-1.015) | .971(.946-.998) |
| Supp Oxygen (Yes) | 2.03 | <.001* | 12.25(5.94-25.25) | 7.66(3.24-18.10) |
| CXR | .461 | .059 | 1.73(1.22-2.46) | 1.585(.981-2.56) |
| Constant | -7.34 |  |  | 0(0-0.01) |

Model1: Variables initially included: Age, gender, CFS, Comorbidities >2, NLR, CRP, creatinine, RR, CPAP, SFR, Total LOS, interaction CPAP\*CFS.
Model2: Variables initially included were the same as model 1+ Supp O2 and CXR.
pseudoR<sup>2</sup><sub>model1</sub> =25.2%, pseudoR<sup>2</sup><sub>model2</sub> =32.6% which mean Sup O2 as a significant predictor could explain additional 7.4% of variance in mortality outcome.
Model1 cross validation accuracy on test set=78.3%, AUROC=.842.
Model2 cross validation accuracy on test set=78.1%, AUROC=.871.

**Figure (1):**
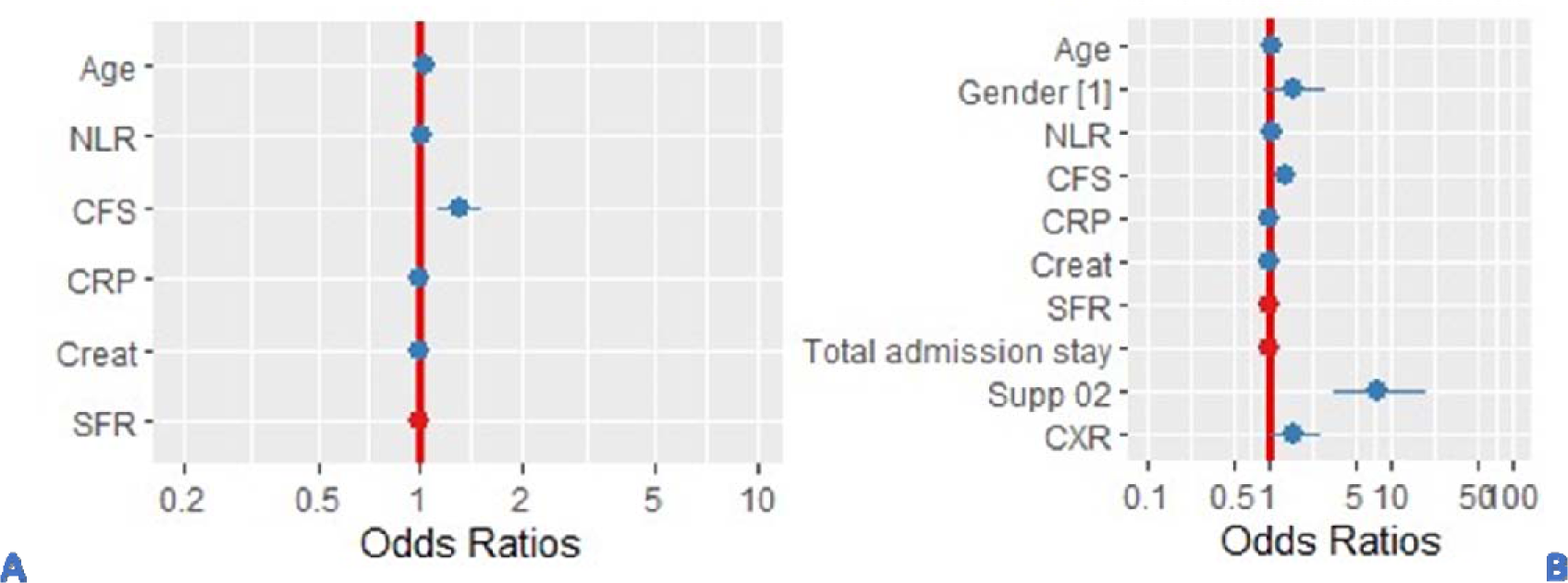
Adjusted Odds ratio calculated from multivariate stepwise logistic regression analysis for assessing independent predictors of mortality. A: model1, B: model2. Likelihood Ratio Test for comparison of goodness of fit between model 1 and 2 revealed better performance in favour of model 2(x^2^=35.8, pc.001*)

## Discussion

This retrospective study identified several risk factors for poor outcomes in hospitalised adults with COVID-19.

The striking observation was the high mortality rate in our cohort, 38% compared to the national average of 33% ^14^. The plausible explanation is that a large proportion of population in Southend are retired and elderly.

The second key finding was older age with greater frailty scores. There are very few studies which evaluated clinical frailty in patients in COVID-19. Similar to our study, an Italian group assessed frailty, which demonstrated increased in-hospital mortality, ICU admissions, independent of age and Sex ^15^. Another study showed that CFS, but not age, remained independently associated with mortality ^16^. Frailty is perhaps a syndrome that is characterised by dysregulation of the innate and adaptive immunity that leads to chronic inflammation, reduced physiologic reserve and increased risk of poor health outcomes. Frailty should be considered in risk assessment models in future studies and clinical trials to assess interventions and meaningful outcomes.

There is substantial literature emphasizing the importance of geriatric medicine toward frailty prevention and clinical criteria to rapidly identify those with frailty or pre-frailty ^17,18^. The irreversible downward spiralling of frailty will begin, if any acute negative health conditions break the equilibrium. This was clearly reflected in the recent COVID -19 pandemic, particularly in countries such as Italy.

In a prospective study of older patients with community acquired pneumonia, nursing home residency was an independent risk factor for viral pneumonia, which highlights the role of frailty in institutionalised populations ^19^ and is associated with worse outcomes in hospitalized older patients *^20,21^*. The UK NICE guidelines recommend the use of CFS in appropriate patients and states that COVID patients with CFS >5, would need to be considered if they were appropriate for critical care management. However, empirical evidence supporting the use of frailty instruments to predict treatment outcomes and triage accordingly is lacking ^22^.

Thirdly, our results confirmed that comorbidities, in particular cardiovascular and diabetes were strongly associated with negative outcomes. This is consistent with recent meta-analysis, from CDC China ^23^. Similarly, another study of 5700 hospitalised patients with COVID-19 in the New York City area, the most common comorbidities were hypertension (57%), obesity (42%), and diabetes (34%)^24^. Of significance, hypertension was reported to increase the odds for death in patients with COVID-19 ^25,26^; however we didn’t find hypertension to be statistically significant. While hypertension does appear to be associated with more severe disease and increased mortality, there is no strong evidence to indicate increased susceptibility of patients with hypertension to COVID-19^27^. The mechanisms of this possible relationship and their clinical relevance have been reviewed in a recent statement of the European Society of Hypertension. The putative relationship between hypertension and COVID-19 may relate to the role of ACE 2 *^27^*. Diabetes, lung diseases, and obesity, are now well recognised major predictors of poor clinical outcomes. These aspects emphasize the importance of the need for multidisciplinary assessment and treatment, including cardiovascular risk evaluation and therapy, during the course of COVID-19 to reduce mortality.

Data shows European mortality is generally higher in older patients compared to earlier reports from China. Age, as an independent predictor of mortality, was observed in our cohort, which was consistent with the large prospective UK ISARIC study of hospitalised patients ^14^ and China ^28-30^. In Italian studies, case fatality rates ranged from 35.5% to 52.5% in patients aged over 70 years with COVID infection ^31-34^. In the USA, older patients aged ≥65 years accounted for higher deaths, with the highest incidence of severe outcomes in patients aged ≥85 years ^35^. Why the disease is particularly dangerous in older people is not yet known and poorly understood at the molecular level. It is clear, however, that advanced age alone is by far the most significant risk factor, independent of underlying comorbidities ^36,37^. An abundance of recent data describing the pathology and molecular changes in COVID-19 patients points to both immunosenescence and inflammaging as major drivers of the high mortality rates in older patients.

In contrast to the literature, male sex was not associated with increased mortality in our study. Large studies from China, Europe and Italy established that males were more susceptible to COVID-19-related complications, representing between 50% and 82% of the hospitalized patients with COVID-19 ^5,8,39^.

We found that baseline CRP, creatinine and NLR were associated with negative impact on mortality. The most consistent prognostic markers in COVID-19 across the different studies were elevated levels of CRP, LDH,Lymphopenia and Neutrophil-to-lymphocyte ratio (NLR), and these appear to stratify patients into higher risk of complications ^40-42^. Intriguingly, elevated levels of C-reactive protein appear to be unique to COVID-19 patients when compared to other viral infections. Other consistently reported markers in non-survivors are increased procalcitonin (PCT) and IL-6 levels ^43^.

### Limitations

The findings of this study are derived from hospitalised cases which might have introduced a bias in disease severity and fatality. The data collection is limited to what is documented in the electronic patient database whether there may be errors both with patient and clinician recall. Our single centre findings may not be generalizable. Routine tests such as LDH, Ferritin, D-Dimer and Troponin could not be carried out on all patients.

### Where do we go from here?

In this large retrospective study, we found that older age, comorbidities, frailty and elevated CRP at admission were significant risk factors for poor outcomes in patients with COVID-19.

Now, more than ever, a holistic approach to patients with comorbidities is required, and rapid solutions to support this must be identified and implemented with urgency. Elderly patients are particularly susceptible to adverse clinical outcomes in COVID -19 infection and assessment and treatment is challenging. Long-stay residential care homes and hospitals need to urgently design adequate health care plans for elderly patients. Our results strengthen the NICE guidance on the Clinical Frailty Scale, to assist decision-making regarding hospitalization. We suggest integrating the frailty assessment in all COVID-19 patients at hospital admission, which can help clinicians in their decision-making processes. However, shared decision-making is always warranted with respect to personal wishes and preferences of the patient. Given the economic and resource constraints, shifting hospice and palliative care resources to the community was a key message in a recent review to inform practice in the pandemic ^44^.

A frailty-based risk-stratification approach, rather than age may prove more valuable when considering interventions in patients with multiple comorbidities. The planning strategies perhaps should include awareness, tools to facilitate communication with healthcare professionals, improved access to institutional health communication and better access to local and social support activities.

## Data Availability

Data is available at reasonable request

## Author Contributions

All authors contributed to data collection, first draft GK, SG and all authors approved final version. Dr GK had full access to all of the data and takes responsibility for the integrity of the data and accuracy of the data analysis. Analysis of the data was performed by Dr Iman El Sayed.

## Conflict of Interest

Dr SG, MD, VW, FH, ID and YA have nothing to disclose. Gouri Koduri reports research support from Roche outside the submitted work.

## Funding

None received

## Notes

### Funding Statement

None recieved

### Author Declarations

This is a retrospective study on COVID Pneumonia which required data collection only. This study was conducted in accordance with the Institutional review board and approved by the Southend University Hospital NHS Foundation Trust and granted a waiver due to its retrospective observational design

